# Biomarker Convert: a novel approach to transform plasma pTau217 results across immunoassay and mass spectrometry analytical platforms

**DOI:** 10.64898/2026.09.28.26363900

**Authors:** Karen J. Quadrini, Jiri Aubrecht, Frederique Vernhes, Xuemei Zeng, Ann D. Johnson, Beth E. Snitz, Ann D. Cohen, Ryan E. Tooker, Mary E. Quiceno, Brian M. Campbell, Thomas K. Karikari, Chad J. Swanson

## Abstract

**INTRODUCTION:** We report the development and validation of a novel computational tool to cross-convert plasma pTau217 results from different assays to facilitate recruitment for therapeutic trials and observational studies.

**METHODS:** Participants underwent Aβ PET imaging and plasma pTau217 measurement on up to three assay platforms across three cohorts. We then developed conversion equations using Deming regression – collectively named *Biomarker Convert* – and evaluated robustness using independent research cohorts. Finally, we modeled the potential impact of pTau217 screening on trial enrollment.

**RESULTS:** While absolute plasma pTau217 concentrations differed, the assays had similar predictive characteristics for Aβ positivity (LC-MS/MS [C_2_N], AUC=0.952; Janssen Simoa, AUC=0.946; Lumipulse, AUC=0.929). *Biomarker Convert* enabled transformation of plasma pTau217 data across assays independent of Aβ PET and clinical status. Plasma pTau217 reduced Aβ PET scans in the model screening paradigm by >60%.

**DISCUSSION:** *Biomarker Convert* equations enable harmonization of plasma pTau217 values from immunoassay and mass spectrometry platforms, supporting direct data comparison.

## 1. Background

Patient selection is critical to the success of clinical trials targeting amyloid beta (Aβ) in Alzheimer’s disease (AD), because efficacy depends on the presence of brain Aβ deposits. Enrollment of patients in large clinical trials evaluating new anti-Aβ therapies is complex, with screen failure rates reaching 80-90% [1, 2]. Effective screening strategies that reduce cost and minimize patient burden are therefore essential.

Inappropriate patient selection has contributed to prior inconclusive anti-Aβ trial outcomes [3, 4]. Aβ PET tracers enable direct *in vivo* visualization and quantification of Aβ plaques and are validated against autopsy findings as a gold standard for detecting AD pathology [5]. However, PET is burdensome and expensive; issues compounded by the fact that PET-related screen failure rates can exceed 40% [6]. CSF biomarkers provide a viable alternative to PET imaging for establishing amyloid pathology. FDA- and EMA-approved biomarkers in CSF, such as Aβ42 (particularly Aβ42/40) and phosphorylated tau fragments, correlate strongly with brain Aβ presence [5]. Nevertheless, CSF sampling is invasive and not uniformly performed worldwide.

Blood-based biomarkers (BBMs) are minimally invasive and can be easily integrated into routine patient laboratory testing with a single blood draw. Plasma biomarkers such as Aβ42/40 ratio, pTau181, and pTau217—including a ratio (or percentage) of its phosphorylated and non-phosphorylated forms—correlate with Aβ plaque status [7]. pTau217 may correlate most strongly with Aβ and tau PET [8], support disease staging [9, 10], and track with CSF levels [11, 12], which reinforces its utility in aiding AD diagnosis. Accordingly, multiple clinical trials have begun including plasma pTau217 as a patient screening tool to help identify Aβ PET positive (Aβ PET+) candidates for anti-Aβ immunotherapy [13, 14]. Because multiple commercial assays are available for pTau217 measurement, methods for comparing results across analytical platforms are critically needed.

Our work builds on prior studies characterizing use of plasma BBMs as a screening tool for individuals with Aβ pathology. We first confirmed that plasma pTau217 outperforms other BBMs for detecting Aβ PET positivity among individuals screened for anti-Aβ immunotherapy trials. Next, we developed a robust method to transform pTau217 results across assay platforms and then modeled whether incorporating plasma pTau217 into a screening cascade could reduce unnecessary Aβ PET imaging.

## 2. Methods

### 2.1. Study Participants

To compare BBM performance to detect Aβ PET positivity, 555 individuals (aged 55 – 85) with gradual and progressive change in memory function for ≥6 months either reported by the subject or study partner or with a diagnosis of Alzheimer’s according to the National Institute on Aging and Alzheimer’s Association (NIA-AA) criteria [15] were evaluated by Aβ PET using either ^18^F-florbetapir (AV45; Amyvid) or ^18^F-florbetaben (BAY 94-9172; Neuraceq) tracers while undergoing screening for phase 1 clinical trials investigating an anti-Aβ immunotherapy, PRX012 [16], from December 2022 to August 2023. Irrespective of the tracer, Aβ positivity was determined by a Centiloid (CL) score of 30 or higher as ascertained by central read [17-19]. *APOE* status was determined using PrecivityAD-APOE (C_2_N Diagnostics, St. Louis, MO) [20]. For individuals who were eligible based on Aβ PET status, a clinical diagnosis of mild cognitive impairment (MCI)/mild AD was provided or confirmed by the clinical site investigator. All individuals consented to study participation using IRB/IEC approved informed consent forms prior to any study procedures.

Of the 555 screened individuals, a representative subset (n=118) screened between January and May 2023 was further evaluated as the exploratory cohort to assess pTau217 performance across analytical platforms. This subset featured an even distribution of Aβ PET+ versus Aβ PET-status across a wide range of CL values (Table 1).

**Table 1.** Demographic Characteristics of the Exploratory and Confirmatory Cohorts.

| Characteristic | Category | Exploratory<br>(N=118) | Confirmatory |  |
| --- | --- | --- | --- | --- |
|  |  |  | MYHAT-NI<br>(N=92) | CoBrA<br>(N=109) |
| Age (years) | n | 118 | 92 | 109 |
|  | Mean (SD) | 70.8 (7.7) | 77.4 (6.4) | 66.5 (9.1) |
| Sex, n (%) | Female | 76 (64.4) | 47 (51.1) | 60 (60.6) |
|  | Male | 42 (35.6) | 45 (48.9) | 43 (39.5) |
| Ethnicity, n (%) | Hispanic or Latino | 66 (55.9) | 0 | 0 |
|  | Not Hispanic or Latino | 52 (44.1) | 88 (95.7) | 62 (56.9) |
| Race, n (%) | Asian | 2 (1.7) | 0 | 1 (0.9) |
|  | Black or African American | 9 (7.6) | 4 (4.4%) | 47 (43.1) |
|  | White | 106 (89.8) | 88 (95.7) | 61 (56.0) |
|  | Not reported | 1 (0.8) | 0 | 0 |
| Height (cm) | n | 45* | 92 | 109 |
|  | Mean (SD) | 165.5 (8.6) | 168.2 (9.4) | 170.5 (10.2) |
| Weight (kg) | n | 45* | 92 | 109 |
|  | Mean (SD) | 70.7 (12.3) | 80.7 (17.3) | 82.6 (15.6) |
| APOE Type, n (%) | E2/E2 | 0 | 0 | 2 (1.8) |
|  | E2/E3 | 5 (4.2) | 9 (9.8) | 14 (12.8) |
|  | E2/E4 | 4 (3.4) | 0 | 5 (4.6) |
|  | E3/E3 | 59 (50.0) | 70 (76.1) | 58 (53.2) |
|  | E3/E4 | 39 (33.1) | 13 (14.1) | 26 (23.9) |
|  | E4/E4 | 11 (9.3) | 0 | 4 (3.7) |
| APOE Category, n (%) | Heterozygous | 43 (36.4) | 13 (14.1) | 31 (28.4) |
|  | Homozygous | 11 (9.3) | 0 | 4 (3.7) |
|  | Non-carrier | 64 (54.2) | 79 (85.6) | 74 (67.9) |

| Characteristic | Category | Exploratory<br>(N=118) | Confirmatory |  |
| --- | --- | --- | --- | --- |
|  |  |  | MYHAT-NI<br>(N=92) | CoBrA<br>(N=109) |
| Amyloid PET (positive)† | n | 59 | 23 | 28 |
|  | Mean (SD) | 76.04 (36.20) | 1.7063 (0.2715) | 1.7469 (0.3146) |
| Amyloid PET (negative)† | n | 59 | 69 | 81 |
|  | Mean (SD) | -4.31 (15.48) | 1.1207 (0.068) | 1.0973 (0.063) |
| LC-MS/MS pTau217 (pg/mL) | n | 118 | 0 | 0 |
|  | Mean (SD) | 1.8592 (1.6344) | NA | NA |
|  | Min, Max | 0.515, 9.303 | NA | NA |
| Lumipulse pTau217 (pg/mL) | n | 116 | 92 | 109 |
|  | Mean (SD) | 0.2765 (0.2783) | 0.1666 (0.1087) | 0.2181 (0.9513) |
|  | Min, Max | 0.0423, 1.602 | 0.0420, 0.7740 | 0.0300, 10‡ |
| Simoa pTau217 (pg/mL) | n | 116 | 92 | 109 |
|  | Mean (SD) | 0.0772 (0.0680) | 0.0594 (0.0309) | 0.0469 (0.0304) |
|  | Min, Max | 0.007, 0.415 | 0.0182, 0.1761 | 0.0111, 0.1789 |

Min, minimum; Max, maximum; n, number of participants; NA, not available
\*Number of participants with available data
†Mean ± SD of Aβ (amyloid) PET results are expressed in Centiloid (CL; exploratory cohort) or Standardized Uptake Value Ratio (SUVR; confirmatory MYHAT-NI and CoBrA cohorts)
‡The second highest value was 0.598 pg/mL

Two confirmatory cohorts, the Monongahela Youghiogheny Healthy Aging Team-Neuroimaging (MYHAT-NI) and Connectomics in Brain Aging and Dementia (CoBrA), were community-based, observational prospective studies composed primarily of cognitively normal participants. MYHAT-NI study aimed to assess the extent of PET-detected AD pathology in dementia-free older adults and explore its associations with sociodemographic and lifestyle factors. It recruited a subset of individuals from its parent MYHAT study, which enrolled participants aged 65 and older from socioeconomically disadvantaged communities in Western Pennsylvania, USA, using age-stratified random sampling from voter records. At enrollment, all MYHAT-NI participants had a Clinical Dementia Rating Global Score (CDR) sum-of-box of less than 1.0. They underwent neuroimaging assessments including ^11^C-PiB PET for Aβ plaques. Additional study design details are provided in previous publications [21-23].

CoBrA aimed at improving the understanding of the relationships between AD and cerebrovascular disease. It recruited participants, aged 50–89, primarily through the University of Pittsburgh Alzheimer’s Disease Research Center and the public Pitt + Me web portal for research volunteers. Each participant underwent extensive neuropsychological assessment including ^11^C-PiB PET for Aβ plaques as previously described [24]. For both MYHAT-NI and CoBrA cohorts, a global ^11^C-PiB standardized uptake value ratio (SUVR) was computed and participants with SUVR >1.346 were classified as Aβ positive (A+) [25-27]. All participants of MYHAT-NI and CoBrA provided written consent, and the University of Pittsburgh Institutional Review Board approved both studies.

### 2.2. Plasma sample collection and biomarker analysis

For the cohort of 555 individuals, whole blood was collected in K_2_EDTA tubes and processed for plasma by centrifugation at 500-700xg for 15 minutes at 4°C. Plasma was aliquoted into standard polypropylene microtubes, frozen on dry ice, and stored at -80°C. Samples were analyzed for Aβ42 and Aβ40 (n=555), pTau181 (n=196), and pTau217 (n=183) using liquid chromatography-tandem mass spectrometry (LC-MS/MS; C_2_N Diagnostics). Matched plasma samples from a subset (n=118) were also analyzed for pTau217 using the Simoa® pTau217 (Janssen) assay on the HD-X automated immunoassay analyzer (Quanterix, Billerica, MA) and the Lumipulse® G pTau 217 plasma assay on the Fujirebio Lumipulse® G1200 fully automated analyzer (Monogram Biosciences, a subsidiary of LabCorp, South San Francisco, CA). For this analysis, 118 individuals had a pTau217 result by LC-MS/MS, and one result from either Lumipulse or Simoa (n=116 for each comparison).

Blood from MYHAT-NI and CoBrA participants was also collected using K_2_EDTA-containing tubes. Plasma was separated by centrifugation at 2000×g for 10 minutes at 4°C, aliquoted into low-bind polypropylene microtubes, and stored at −80°C. On the day of analysis, samples were thawed at room temperature, vortexed, and then centrifuged at 4000xg for 10 min at 4°C to remove particulates. The Simoa® pTau217 (Janssen) assay was carried out on the HD-X analyzer and the Lumipulse® G pTau 217 Plasma assay on the Fujirebio Lumipulse® G1200 analyzer, similar to the exploratory cohort. All measurements were conducted in the Biomarker Biofluid Laboratory, Department of Psychiatry, School of Medicine at the University of Pittsburgh.

### 2.3. Statistical analysis

Receiver operating characteristic (ROC) area under the curve (AUC) analyses were used to evaluate biomarker ability to differentiate individuals with Aβ load ≥ 30 CL per Aβ PET. Correlation of Aβ PET with plasma biomarkers was evaluated using Spearman’s rank correlation (SAS version 9.4; SAS Statistical Institute, Cary, NC). Differences in each ROC AUC relative to Aβ42/40 AUC were calculated using the DeLong method [28]. The relationships of pTau217 concentrations measured by the three assay platforms were evaluated on log_10_ transformed data using Deming regression in SAS version 9.4 and R version 4.5.2 (R Foundation, www.r-project.org). MYHAT-NI and CoBrA analyses – pTau217 measurements, Deming regression, and conversion equation validation – were performed using MATLAB version R2024a (MathWorks, Natick, MA). Additional statistical analysis was performed using GraphPad Prism version 10.6.1 (GraphPad, San Diego, CA). To enable conversion of log_10_ transformed pTau217 results across assay platforms, *Biomarker Convert* equations were derived from Deming regressions performed using the matched biomarker values for each platform comparison [29].

## 3. Results

### 3.1 Performance of BBMs to detect brain amyloid beta positivity

In an initial assessment, we retrospectively evaluated plasma levels of Aβ42/40, pTau181 and pTau217 in a cohort of screened individuals (up to n=555) using mass spectrometry assays. These biomarkers were assessed for their ability to predict Aβ PET positivity using ROC analysis with 30 Centiloids (CL) as the threshold for Aβ PET positivity [17] (Figure 1). This cohort featured a 30% prevalence of Aβ PET+ participants (167 Aβ PET+ and 388 Aβ PET negative [Aβ PET-]) covering a wide range of PET CL values (-59.19 to 170.99 CL).

**Figure 1.**
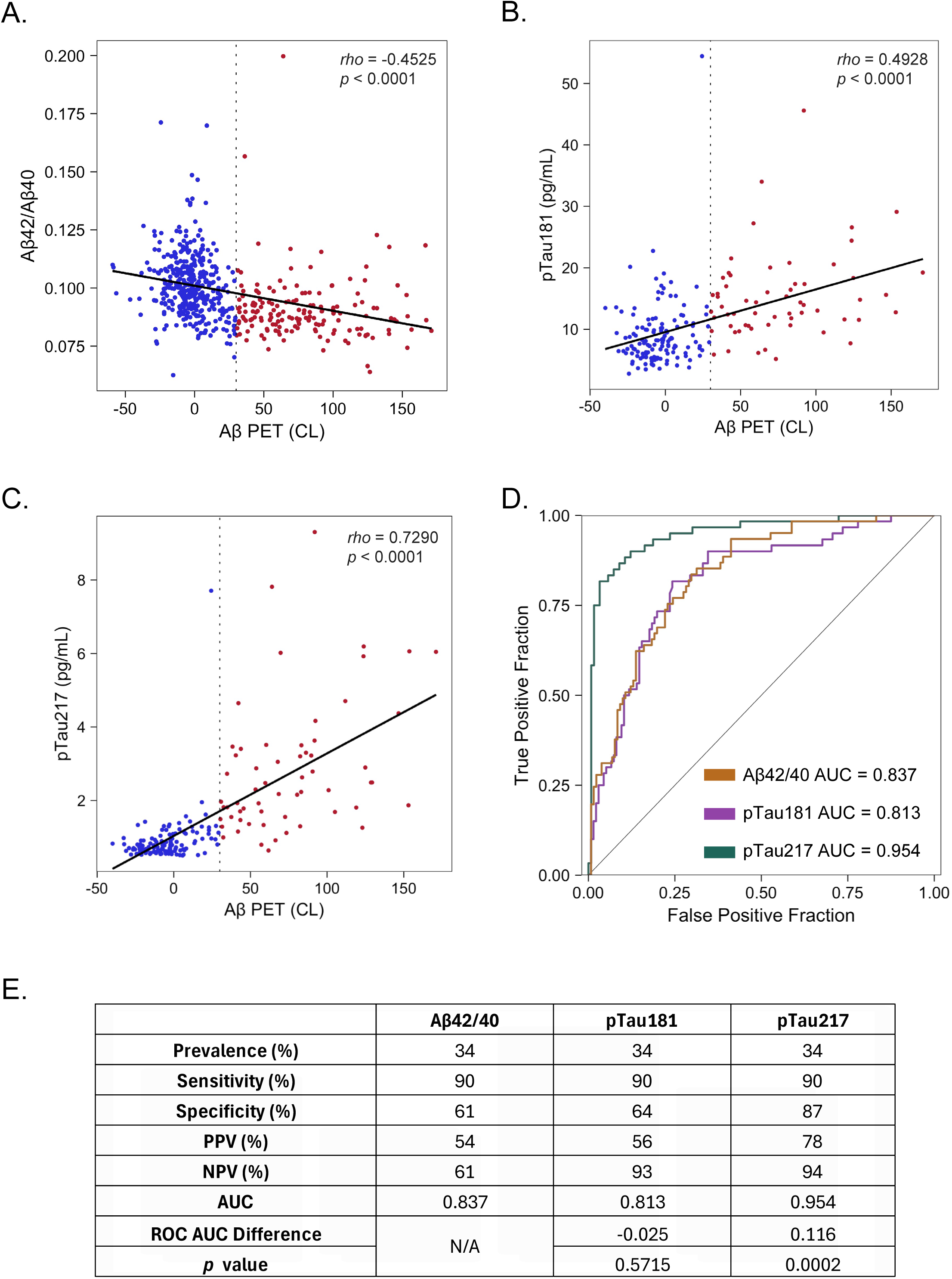
Performance of select plasma biomarkers to detect Aβ PET positivity. Scatterplots of Aβ PET load (≥30 Centiloid cutoff for positivity; vertical dotted line) and plasma Aβ42/40 ratio (n=555) **(A)**, pTau181 concentration (n=196) **(B)**, and pTau217 concentration (n=183) **(C)** each assessed by LC-MS/MS. Individual Aβ PET status is indicated by color (positive = red; negative = blue). Spearman’s *rho* correlation coefficient is noted for each comparison. **D)** ROC curves for plasma pTau181 and pTau217 concentrations and the Aβ42/40 ratio. This analysis was based on a complete dataset for all biomarkers available for direct comparison with Aβ PET status (n=174). **E)** Specificity, sensitivity, and positive and negative predictive values (PPV, NPV) of each biomarker using a 90% sensitivity threshold in this population (n=174) with 34% prevalence of Aβ PET positivity. ROC AUC differences are calculated relative to Aβ42/40, with *p* values indicating the statistical significance of the differences. CL, Centiloid; N/A, not applicable.

As expected, plasma Aβ42/40 ratios had an inverse correlation with Aβ PET CL values whereas pTau181 and pTau217 concentrations were positively correlated with brain Aβ load (Figure 1A-C). All three biomarkers demonstrated good performance at distinguishing Aβ PET positive from negative individuals in a direct comparison in the subset of individuals having results for all three biomarkers (n=174) (Figure 1D). Plasma pTau217 differentiated Aβ PET+ subjects with the highest accuracy (AUC=0.95), whereas the performances of pTau181 and the Aβ42/40 ratio were lower (AUC = 0.81 and 0.84, respectively).

In this matched population, there was an observed prevalence of 34% Aβ PET positivity (59 Aβ PET+ and 115 Aβ PET-). When sensitivity was fixed at 90% for use as a screening tool in a clinical trial, consistent with recommended AD diagnostic criteria [30, 31], specificity was lowest for Aβ42/40 (61%) and highest for pTau217 (87%) (Figure 1E).

### 3.2 Exploratory subset and confirmatory cohort demographics and characteristics

For the plasma pTau217 cross-platform assessment and conversion equation generation, we used a subset of 118 individuals covering a broad range of Aβ PET CL values. This cohort consisted of individuals who reported gradual, progressive change in memory function, of whom 48 were subsequently confirmed as cognitively impaired (40.7%). To evaluate the robustness of the *Biomarker Convert* equations between the Lumipulse and Simoa assays, we used two independent community-based confirmatory cohorts, MYHAT-NI and CoBrA. For the MYHAT-NI cohort, 8 out of 92 (8.7%) were classified with mild cognitive impairment based on a CDR sum-of-box and global score of 0.5. The proportion of individuals clinically diagnosed with cognitive impairment (either MCI or AD) in the CoBrA cohort was 32 out of 109 (29.6%) at the time of enrollment. Available shared characteristics for participants in the three cohorts are presented in Table 1.

Participants in the exploratory cohort subset at baseline showed equal distribution (50%) of Aβ PET positive and negative subjects (59 Aβ PET+, n=118) (Figure 2A). The proportion of Aβ PET+ participants was lower at approximately 25% for each of the confirmatory cohorts, MYHAT-NI (23 Aβ PET+, n=92) and CoBrA (28 Aβ PET+, n=109). The distribution of pTau217 concentrations within these three independent cohorts as measured using two immunoassay platforms varied, with the widest range of concentrations observed in the exploratory cohort (Figure 2B and C). The exception was an outlier in the CoBrA cohort for an Aβ PET-participant with a pTau217 concentration measuring at 10 pg/mL (Table 1).

**Figure 2.**
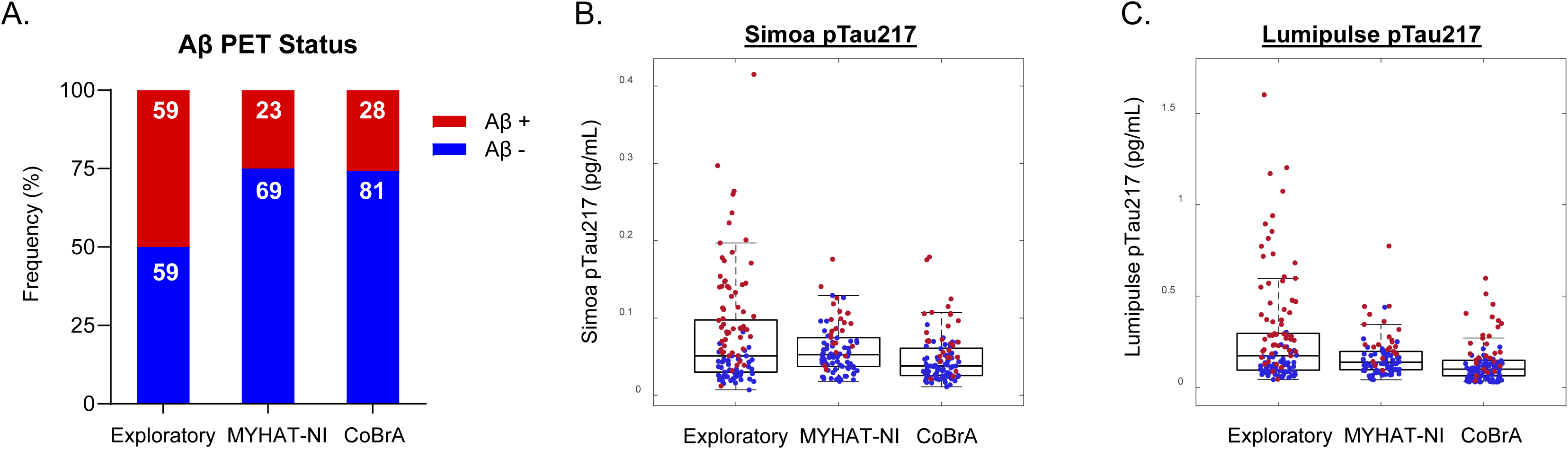
Distribution of biomarker characteristics for the exploratory and confirmatory cohorts. **A)** Aβ PET status is depicted as the frequency of individuals within the cohort (%). The sample size for each subgroup is indicated numerically within the bars. **B)** and **C)** The box plots report the median raw absolute pTau217 concentrations and the range of lower and upper quartiles obtained for each cohort from the **B)** Janssen Simoa and **C)** Lumipulse immunoassays (outlier excluded). The whiskers extend to the most extreme data points within 1.5 times the interquartile range (IQR) from the quartiles. Aβ PET positive = red; Aβ PET negative = blue.

### 3.3 Head-to-head pTau217 cross-platform comparison

Since pTau217 has emerged as the most promising BBM for detecting brain Aβ pathology, several assays with different analytical characteristics have been developed for its measurement. Therefore, we investigated the impact of the analytical method on pTau217 performance in identifying Aβ PET+ individuals (brain Aβ load ≥30 CL). For this assessment, alongside LC-MS/MS, we selected two widely used immunoassays—the Janssen Simoa and Lumipulse pTau217 assays—based in part on their routine use in clinical (Lumipulse) or drug development and clinical research (Janssen Simoa) laboratories. We evaluated matched samples from a representative subset of the screened individuals from the initial cohort who exhibited a wide range of pTau217 levels and where Aβ PET status information (positive or negative) was available (n=118). Our experimental setup followed CLSI guidelines for assay comparison [29], using Deming regression per recommendations for method comparison and bias estimation [32].

Absolute concentrations of plasma pTau217 differed across assays (Table 1). LC-MS/MS produced the highest absolute values with a mean ± SD of 1.8592 ± 1.6344 pg/mL (n=118), while Lumipulse results were approximately 7-fold lower (mean ± SD: 0.2765 ± 0.2783 pg/mL, n=116). The Janssen Simoa assay provided the lowest values, approximately 24-fold lower than LC-MS/MS (mean ± SD: 0.077 ± 0.0680 pg/mL, n=116). The magnitude of difference prohibited direct comparisons of results from the analytical methods.

Despite these differences in pTau217 absolute concentrations, there was a strong linear relationship between the mass spectrometry and immunoassay platforms (Figure 3A-C). Moreover, all three assays accurately detected Aβ positivity with AUC values ranging from 0.929 to 0.952 (Figure 3D). Given this high concordance, we developed a set of equations for the conversion of values to enable platform comparison of pTau217 readouts that we named *Biomarker Convert* (Figure 3E).

**Figure 3.**
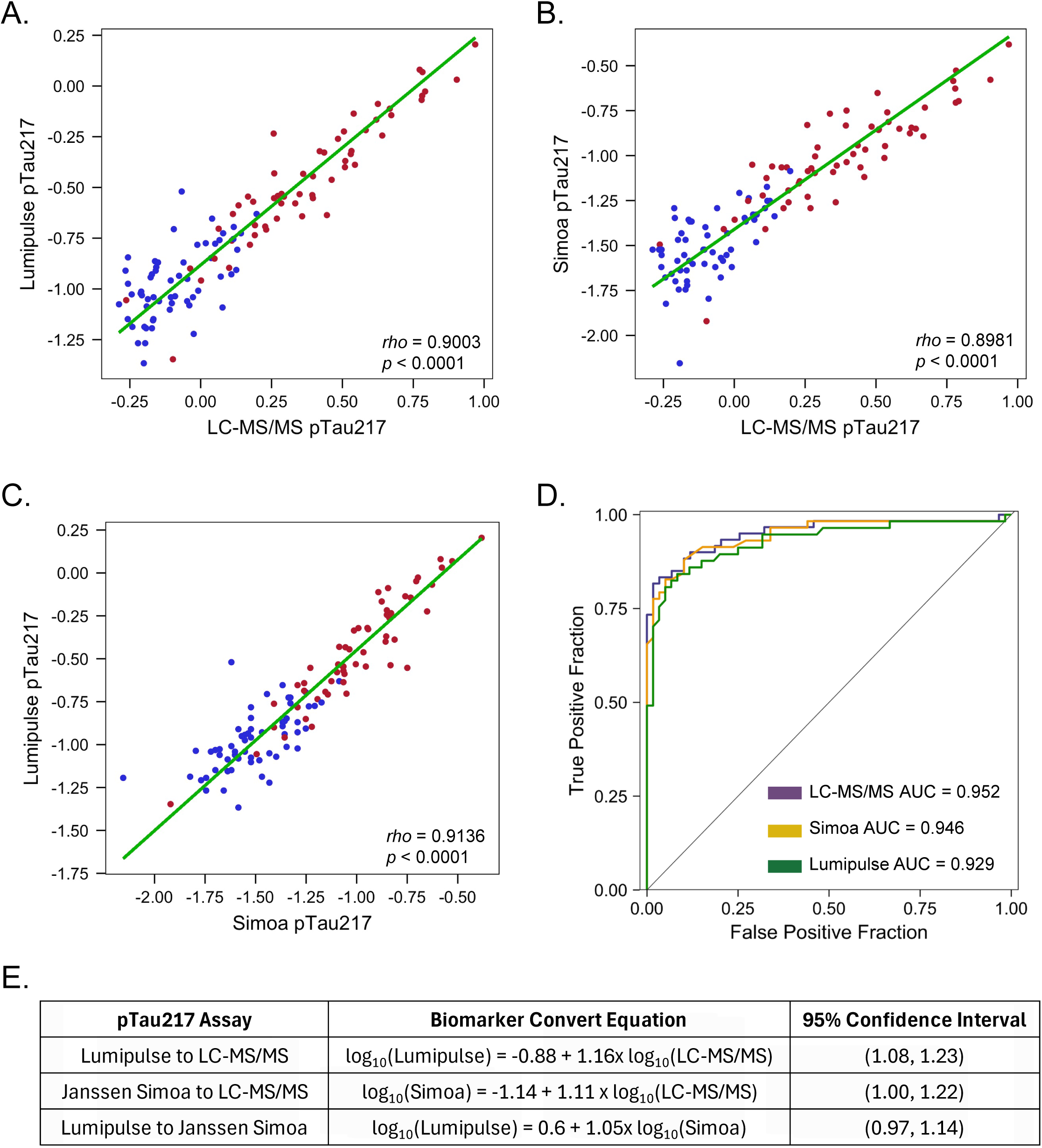
Comparability of pTau217 results across mass spectrometry and immunoassay platforms. **A**-**C)** Matched pTau217 samples were evaluated using LC-MS/MS and two immunoassay platforms, Janssen Simoa and Lumipulse. Log transformed results are presented for pTau217 values. Comparison of LC-MS/MS to either Lumipulse or Simoa (n=116) **(A**, **B)**, and Lumipulse to Simoa (n=114) **(C)** indicated strong direct correlation between pTau217 concentration and Aβ PET positivity regardless of analytical platform (Spearman’s *rho*). Aβ PET status is indicated for each participant (positive = red; negative = blue). **D)** ROC analyses of pTau217 values from each platform to predict Aβ positivity. **E)** *Biomarker Convert* equations generated from Deming regression of the matched pTau217 assay results with 95% confidence intervals indicated for the slopes.

### 3.4 Evaluation of utility of Biomarker Convert equations using independent cohorts

To assess the robustness and general applicability of the *Biomarker Convert* equations for pTau217 assays, we utilized MYHAT-NI and CoBrA cohorts. In contrast to the exploratory cohort, these confirmatory cohorts featured a higher proportion of individuals who were Aβ PET-. Selection of cohorts that comprised different proportions of individuals with distinct baseline characteristics (e.g., predominantly cognitively normal cohorts) allowed us to discern the impact of clinical status on the ability of the *Biomarker Convert* equations to harmonize pTau217 measurements. Samples from these confirmatory cohorts were analyzed independently using the Lumipulse and Janssen Simoa assays (Figure 2B and C).

First, pTau217 results were compared to each other to assess intra-laboratory comparability (self-trained). These intra-laboratory cross-platform pTau217 results also demonstrated strong correlation between the two assay platforms (Figure 4A and B). Next, *Biomarker Convert* equation utility was assessed. For the MYHAT-NI cohort, the median difference in raw absolute values between the assay platforms was -0.416 (Figure 4C). Application of the corresponding *Biomarker Convert* equation reduced the median difference to 0.118, similar to the difference after application of an intra-cohort trained Deming regression.

**Figure 4.**
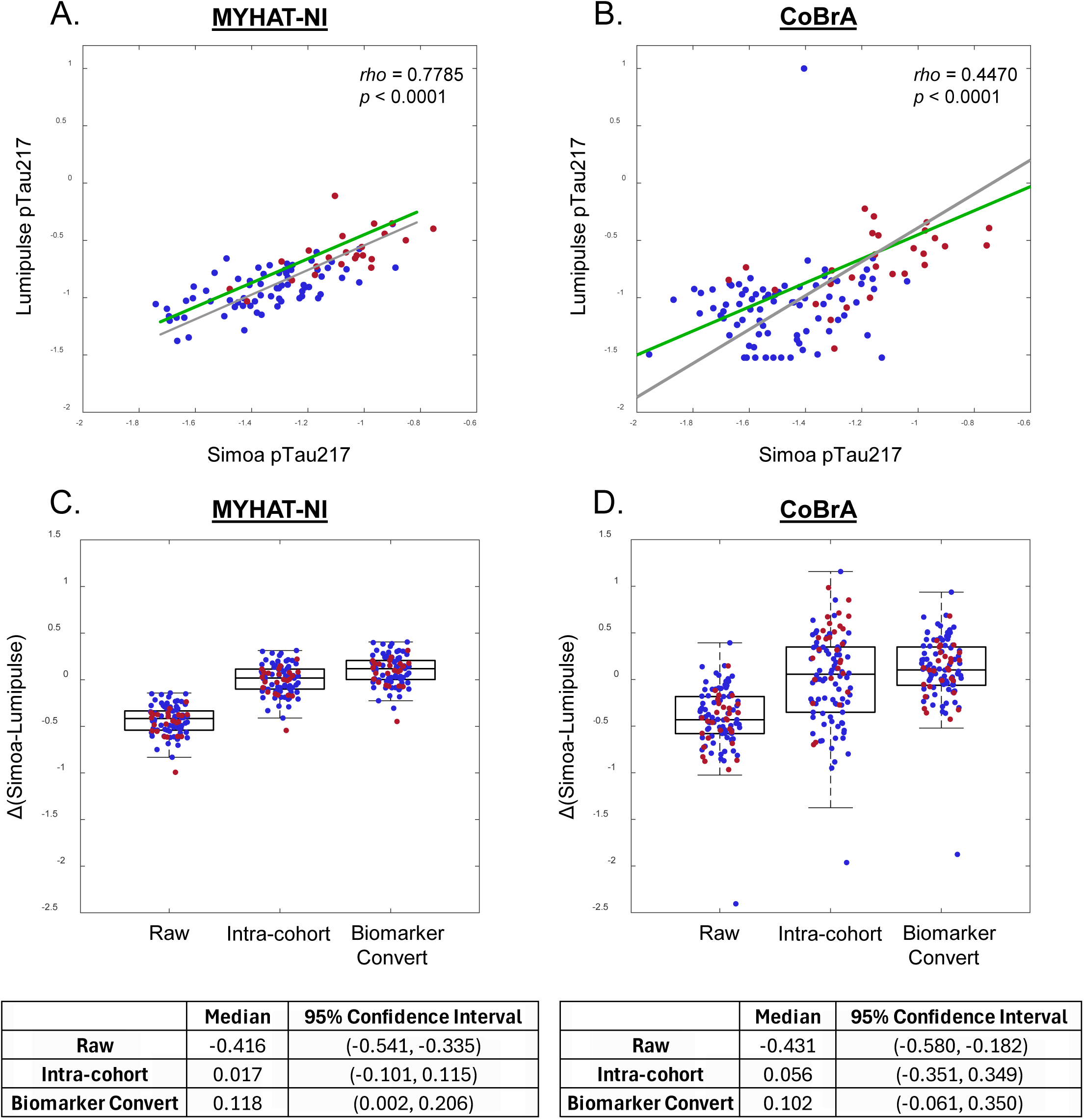
Comparability of pTau217 results across confirmatory MYHAT-NI and CoBrA cohorts. Correlation of Lumipulse and Janssen Simoa pTau217 results (log transformed) in **A)** MYHAT-NI and **B)** CoBrA cohorts. The gray regression line and Spearman’s *rho* correlation coefficient were calculated for the respective confirmatory cohort datasets. The green line represents the regression calculated from the exploratory cohort dataset. The utility of the *Biomarker Convert* equations was assessed in the **C)** MYHAT-NI and **D)** CoBrA cohorts by comparing the differences in raw absolute values, the values calculated using an intra-cohort derived equation, and the values calculated using the assay-appropriate *Biomarker Convert* equation. Box plots report the median difference and ranges of lower and upper quartiles. The whiskers extend to the most extreme data points within 1.5 times the interquartile range (IQR) from the quartiles. The cohort median and 95% confidence intervals are tabulated. Aβ PET positive = red; Aβ PET negative = blue.

For the second confirmatory cohort, CoBrA, the *Biomarker Convert* equation reduced the median difference in pTau217 results from the original -0.431 to 0.102, compared with 0.056 after intra-cohort correction (Figure 4D). Altogether, these observations, using two independent confirmatory cohorts, suggest that the appropriate *Biomarker Convert* equation can normalize pTau217 measurements generated at independent laboratories with different bioanalytical platforms and patient / study populations.

### 3.5 Impact of plasma pTau217 on screening participants for confirmatory Aβ PET imaging in clinical trials

We evaluated the impact of plasma pTau217 screening on selection of brain Aβ positive subjects for clinical trials using parameters from a model clinical study screening process (Figure 5). In the traditional screening paradigm, we conservatively estimated an Aβ PET positivity prevalence of 25% for our modeled process (observed prevalence of 34%; Figure 1E). Under this assumption, approximately 11% more individuals would need to be screened to obtain the same number of eligible, enrolled participants (n=100). However, including this screening step prior to Aβ PET analysis would also substantially reduce the total number of Aβ PET scans performed. Specifically, 427 of 637 (approximately 67%) screened individuals would fail eligibility criteria based on plasma pTau217 levels that would not meet the designated cutoff (defined by 90% sensitivity for this context of use) before undergoing Aβ PET assessment. As a result, implementing plasma pTau217 would decrease Aβ PET examinations by more than 60% in this model.

**Figure 5.**
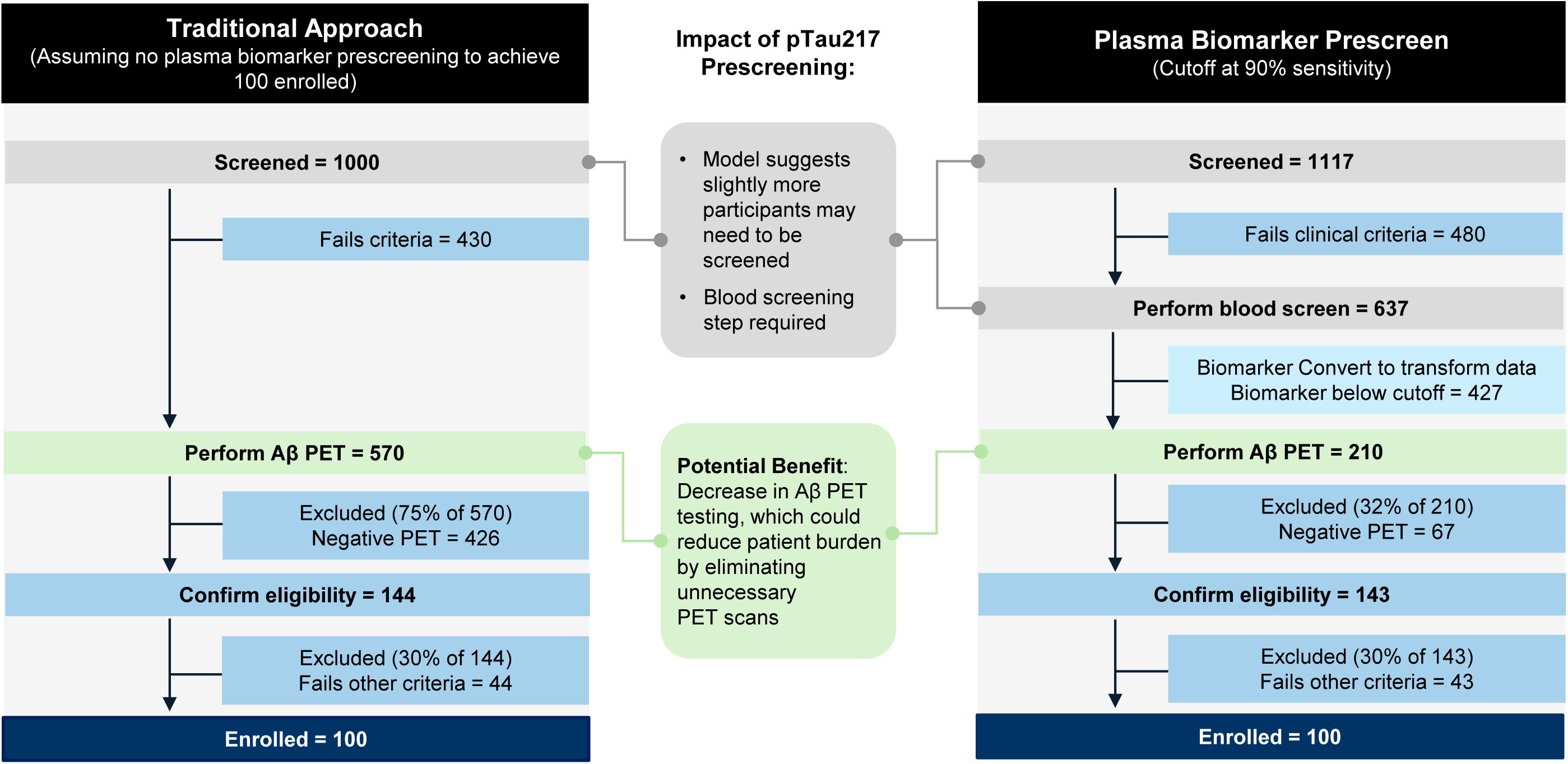
Model for estimating the impact of implementing plasma pTau217 as a screening biomarker during clinical trial enrollment. Clinical trial enrollment approaches depicting use of Aβ PET imaging as a biomarker assessment for eligibility determination (traditional approach; left) versus a two-tiered approach where plasma pTau217 serves as a screening biomarker test prior to confirmatory Aβ PET imaging (pTau217 prescreen approach; right). The number of individuals and percentage that fail defined eligibility criteria or progress from one step to the next are depicted for each approach. *Biomarker Convert* equations could be applied to convert plasma pTau217 measurements from any of three analytical platforms to determine whether the biomarker concentration falls above/below the established cutoff (see Discussion).

## 4. Discussion

Appropriate patient selection is essential for developing anti-Aβ therapies [2, 6, 33]. Clinical symptom-based enrollment has historically produced heterogeneous populations with varying Aβ burden, including participants without Aβ pathology [4, 34, 35]. Yet gold standard Aβ PET increases burden, scheduling bottlenecks, logistics complexity, timelines and cost. CSF Aβ42/40 and pTau isoforms provide alternatives to PET imaging but require lumbar puncture and are not universally adopted. Therefore, more accessible precision tools could increase the likelihood of treatment response while minimizing development risk.

BBMs offer minimally invasive, faster, and less costly detection of brain Aβ pathology. In recognition of the growing clinical utility of BBMs, the FDA granted 510(k) clearance in May 2025 to the first plasma pTau217 assay—Fujirebio’s Lumipulse® G pTau217/Aβ1-42 plasma ratio—as an in vitro diagnostic test for confirming AD [36]; the assay subsequently received an EU CE mark in May 2026 [37]. Since then, additional BBM assays have received FDA clearance or received an EU CE mark including Roche’s Elecsys pTau181 test, Roche’s Elecsys pTau217 test and C_2_N Diagnostics’ PrecivityAD2. In drug development, BBMs have been used for initial screening in several clinical trials generally as laboratory derived tests under CLIA regulations. For example, elevated plasma pTau217 was an inclusion criterion for Trailblazer-ALZ 3 [14] and in conjunction with plasma Aβ42/40 for the AHEAD 3-45 study in preclinical AD [13, 38].

In this study, we compared the performance of three well-studied AD plasma biomarkers, i.e. Aβ42/40 ratio, pTau181 and pTau217, to identify subjects with brain Aβ pathology (≥30 CL), a threshold that limits false positives [39], in a cohort of subjects screened for clinical trial inclusion. All plasma biomarkers correlated with Aβ positivity, with pTau217 outperforming pTau181 and Aβ42/40 ratio consistent with prior studies [8, 40, 41]. Although criteria for Aβ PET positivity differs between studies, the plasma pTau217 AUC (0.95) achieved here agreed with previous reports [40-43], further supporting its robustness and reproducibility across cohorts and analytical assay platforms.

Several plasma pTau217 assay platforms including mass spectrometry and immunoassays exist, so we evaluated the performance and comparability of three that are commonly used. All performed almost identically to detect Aβ positivity, with the mass spectrometry assay having slightly higher AUC than the two immunoassays. These results demonstrate comparable performance among assays despite different reagents and technologies, and are consistent with reports that mass spectrometry may provide higher diagnostic accuracy [8, 43-45]. Despite the small differences in the performance of the assays, ours and other published data suggest that they could be used interchangeably for detecting Aβ positivity [12, 46]. Importantly, given the relatively small differences in diagnostic performance, assay selection to support drug development should be based on operational characteristics of the assay and availability.

Absolute pTau217 concentrations differed across assay platforms, consistent with previously reported results [8, 43, 46-48]. Given these varied measurements, it is essential to develop and apply assay-specific cutoff values to interpret concentration data for determination of Aβ positivity. However, differences in absolute concentration values hinder direct data comparability, complicating interpretation in clinical trials where multiple assays may be used within the same study, for instance, for screening and patient monitoring due to operational considerations. Therefore, the development of approaches enabling seamless comparison of data across assays is essential. *Biomarker Convert* equations provide an initial step for harmonizing measurements outside of efforts to identify certified reference materials [48].

Our *Biomarker Convert* equations were developed in accordance with CLSI principles [29], using samples spanning a wide range of pTau217 concentrations, and the robustness and general applicability of these equations for the Simoa and Lumipulse assays were evaluated using two independent confirmatory cohorts. In contrast to the deliberately balanced samples used for the development of the equations, these two confirmatory cohorts were comprised of predominantly Aβ PET negative cognitively unimpaired individuals. Our equation normalized the pTau217 values, demonstrating that these equations are not affected by differences in bioanalytical laboratories or cohort characteristics such as cognitive status. While additional validation is needed, our data indicates that the *Biomarker Convert* equations can be useful for laboratories, investigators, or groups lacking sufficiently large sample populations to derive their own equations.

Enrollment of participants with Aβ pathology for anti-Aβ clinical trials is a complex endeavor that requires costly Aβ PET imaging and causes significant burden to patients and caregivers. Our model suggests that plasma pTau217 prescreening could reduce the need for eligibility-related Aβ PET scans by >60%, lowering costs and easing patient and clinical site burden. Our analysis complements modeled results showing significantly reduced Aβ PET or CSF testing when plasma pTau217 prescreening for Aβ pathology is implemented in a two-tiered screening approach for asymptomatic preclinical AD trials [45]. However, eligibility criteria and the specific clinical trial population may impact the ultimate reduction rate observed. In a two-tiered screening paradigm, we suggest that the *Biomarker Convert* equations could be used as a general analytical tool to convert pTau217 results from different analytical platforms into values for a single platform with an established cutoff, followed by confirmatory Aβ PET imaging.

There are a few limitations to our study. We could not independently assess the conversion equations between mass spectrometry and the immunoassay platforms due to a lack of matched values in the validation cohorts. Given the high concordance between the various assays, we expect these equations would enable data transformation between LC-MS/MS and individual immunoassays, but confirmation with matched results is needed. Furthermore, testing additional independent cohorts including Aβ positive individuals with higher plasma pTau217 values will be essential to validate the utility of the *Biomarker Convert* equations in this population. Further development could support cross-cohort comparisons and integration of historical, community-based, and clinical trial datasets.

In conclusion, Aβ PET imaging or CSF confirmation of AD pathology is currently the gold standard for diagnosis and clinical trial enrollment. As plasma pTau217 gains greater acceptance and use as a diagnostic biomarker, it may reduce reliance on these invasive and expensive tests, but multiple commercial assays limit data comparability. Consequently, harmonization methods or reagent standardization is essential for routine use in clinical trials and clinical practice. The presented *Biomarker Convert* equations provide an approach for harmonizing plasma pTau217 readouts across analytical platforms and provides a practical solution to support wider implementation of plasma pTau217 in future clinical trials.

## Acknowledgments

The authors thank all the study participants and their care partners, the study site staff, and investigators for their time and commitment. The authors would also like to thank Kevin Dougherty and Courtney Fitzgerald for their operational assistance during this study; and Allyson O’Neil and Meenakshi Khare (Quanterix), Tim Persyn (Monogram Biosciences) and John Bui (C_2_N Diagnostics) for data management and technical assistance.

## Author Contributions

JA and KJQ conceptualized the project and supervised the study. ADJ, FV and XZ performed statistical analysis. BES and ADC led cohort recruitment for the MYHAT-NI and CoBrA studies. KJQ, JA, FV, ADJ, XZ, and TKK interpreted data. KJQ authored the initial draft of the manuscript and oversaw manuscript preparation. CJS, MEQ, RET, and BMC provided intellectual contributions and reviewed and edited the manuscript. KQ, JA, BMC and TKK wrote the final manuscript. All authors reviewed and approved the final manuscript.

## Conflict of Interest Statement

KJQ, ADJ, BMC, and CJS are employees of Prothena Biosciences Inc and stockholders of Prothena Corporation plc; JA, FV, RET and MEQ were employees of Prothena Biosciences Inc at the time of these studies; KJQ, JA, FV, BMC and CJS are inventors on Prothena patents or patent applications. XZ is an employee of the University of Pittsburgh Medical Center; TKK, BES and ADC are employees of University of Pittsburgh School of Medicine and, over the last two years, has served as an ad hoc consultant and/or advisory board member for Quanterix Corporation, Spear Bio Inc., Neurogen Biomarking LLC., Alzheon, Siemens Healthineers and Neurogen Biomarking LLC, and has received honorarium from Cell Signalling Technologies. TKK and XZ are inventors on patents and provisional patents regarding biofluid biomarker methods, targets and reagents/compositions, that may generate income for the institution and/or self should they be licensed and/or transferred to another organization. TKK has received royalties from Bioventix for the transfer of specific antibodies and blood biomarker assays to third-party organizations. The remaining authors declare no competing interests.

## Funding Statement

This study was sponsored by Othair Prothena Limited, Dublin, Ireland, a member of the Prothena Corporation plc group. BES and the MYHAT-NI study are supported by the NIA (R01 AG052521 and R01 AG023651). TKK and the Karikari Laboratory are supported by the National Institutes of Health (NIH) (R01 AG083874, U24 AG082930, P30 AG066468, RF1 AG052525, R01 AG053952, R37 AG023651, RF1 AG025516, R01 AG073267, R01 AG075336, R01 AG072641, P01 AG025204) and a professorial endowment from the Department of Psychiatry, University of Pittsburgh. No other funding supported this study.

## Consent Statement

All participants provided written informed consent prior to any study procedures. All individuals in the exploratory cohort [n=555] were consented by Advarra central IRB or IEC approved informed consent forms prior to study participation. All participants of MYHAT-NI and CoBrA were consented by the University of Pittsburgh Institutional Review Board.

## Data availability statement

On request, and subject to certain criteria, conditions, exceptions and applicable data privacy laws, Prothena will provide access to deidentified participant data from the applicable Prothena-sponsored cohort. Aggregated data are available for the University of Pittsburgh cohorts from TKK on reasonable request.

## Notes

### Author Declarations

This study was conducted in accordance with all relevant ethical guidelines. Ethical approval for the exploratory cohort was granted by the Advarra Institutional Review Board (IRB) in accordance with US FDA regulations (21 CFR Parts 50 & 56). US participants were consented by Advarra IRB/Independent Ethics Committee (IEC) approved consent forms. Local approvals in Spain and Poland were obtained from the CEIC Grupo Hospitalario Quirón Barcelona and Komisja Bioetyczna przy Bydgoskiej Izbie Lekarskiej, respectively. The University of Pittsburgh IRB approved the MYHAT-NI (STUDY19020264) and CoBrA (STUDY19100015C) cohorts. All participants provided written informed consent prior to study procedures.

